# What should be included in an educational programme for non-health professional exercise instructors in charge of community-based exercise targeting young adults in antipsychotic treatment – A focus group study of stakeholder perspectives

**DOI:** 10.1101/2023.08.03.23293602

**Authors:** MF Andersen, K Roed, V Sørensen, A Riis, BS Rafn, BH Ebdrup, J Midtgaard

## Abstract

**Background:** Exercise plays a crucial role in addressing the increased cardiometabolic morbidity and premature mortality in people with schizophrenia. When delivered in community-based settings, exercise may also reduce loneliness, while promoting overall physical active behaviours. Skilled instructors are essential to deliver effective community-based exercise; however, knowledge about their roles and required training is lacking. We investigated stakeholders’ perspectives on components needed for an educational programme for non-health professional exercise instructors delivering community-based exercise targeting young adults in antipsychotic treatment.

**Methods:** We conducted six focus groups comprising a total of 30 individuals representing five different stakeholder groups, namely clinical staff within mental health, physiotherapists, exercise instructors, young adults in antipsychotic treatment, and relatives to young adults in antipsychotic treatment. Data were analysed using qualitative content analysis, as described by Graneheim and Lundman.

**Results:** The analysis identified three categories: (i) *acknowledging mental illness*, (ii) *applying a resource-oriented approach*, and (iii) *promoting exercise as a shared activity*, and one overarching theme: *instructors as guardians of an inclusive culture*.

**Conclusions:** An educational programme for exercise instructors delivering community-based exercise to young adults in antipsychotic treatment should focus on securing an inclusive culture that embraces an anti-stigmatising approach. Results of the current study informed the development of an educational programme consisting of an instructor manual, a one-day educational programme for instructors, and a continuous exchange of experiences between instructors.

## Background

Increasing evidence suggests that physical activity encompassing exercise and sports plays a crucial role in addressing the increasing prevalence of comorbidity, e.g., diabetes (1), metabolic syndrome, cardiovascular disease (2), and premature mortality (3–5) in people with schizophrenia. Also, exercise can facilitate the long-term clinical recovery process in people in the early stages of schizophrenia (6). Specifically, increasing evidence suggests that community-based sports and exercise in groups can reduce social isolation and stigmatisation, as well as enhance the social identity of people with schizophrenia (7–9). Furthermore, community-based sports might serve as an opportunity to provide life skill training, improve social connectivity, and promote overall physically active behaviours in the same intervention (10). Gym-based group exercise is both feasible (11) and more popular among people with psychosis compared to other sporting activities (12). Still, people with schizophrenia engage in less physical activity (13) and have more sedentary behaviour (14) compared to the general population. Early intervention is critical for the optimal treatment of schizophrenia (15,16), and the early stages of the illness may constitute a window of opportunity to establish sustainable physical activity habits when patients are younger, more active, and less affected by physical comorbidities (17,18). Consequently, building sustainable, easily accessible, and engaging community-based exercise and sporting activities targeting people with schizophrenia has gained increased attention among clinicians, politicians, and stakeholders.

Qualitative studies have indicated that, for people with severe mental illness, simply moving from intention to participation is insufficient to secure engagement in community-based exercise in groups. Long-term engagement requires intensive practical and emotional support (9,19), and here instructors delivering and supervising community-based exercise play a key role (20). The mental health literature widely recognises that exercise and lifestyle interventions supervised by professionals with relevant training, such as physical educators, physiotherapists, and exercise physiologists are associated with greater improvements compared to unsupervised interventions and/or interventions supervised by other health professionals (21). We recently investigated the perspectives of professional experts in relation to developing community-based exercise programs for people with schizophrenia. The results of this study (*Andersen et al., submitted*). emphasized the importance of ensuring safe and meaningful exercise content and that exercise instructors receive formal education prior to facilitating community-based exercise for people with schizophrenia. However, the specific elements of the role and the required skills of exercise instructors have attracted little research attention, and accordingly, the content of an educational programme remains unexplored.

Thus, the aim of the current study is to investigate the perspectives of various stakeholders in relation to the development of an educational programme for non-health professional exercise instructors delivering community-based exercise for young adults in antipsychotic treatment.

## Methods

Design and setting

The study was conducted as part of the development and design of the Vega trial (ClinicalTrials.gov identifier: NCT05461885), which aims to evaluate the effectiveness of a gym-based exercise programme delivered by non-health professional exercise instructors for young adults receiving antipsychotic medication. The current study applied a descriptive qualitative design using researcher triangulation and semi-structured focus groups. The study was reported according to the Consolidated Criteria for Reporting Qualitative Research checklist (22) (Appendix 1).

Sampling

We used a purposeful sampling strategy to ensure information richness (23), i.e., by recruiting participants with specific knowledge, experience, and/or interest concerning the study aim.

Specifically, stakeholders targeted or affected by a community-based exercise intervention were considered relevant contributors of important knowledge on the required training of exercise instructors (24). Thus, we recruited representatives from the following five main stakeholder groups: clinical staff from outpatient mental health services; physiotherapists working in mental health care; exercise instructors, both with and without previous experience with facilitating exercise for young adults in antipsychotic treatment; young adults in antipsychotic treatment; and relatives of young adults in antipsychotic treatment. Within the stakeholder groups, we recruited using convenience sampling through email invitations distributed by gatekeepers to the different stakeholder groups, e.g., the team leader of clinical staff, peer-board coordinator, or personal email directly to stakeholders such as exercise instructors. We aimed to conduct focus groups comprising four to seven stakeholders, as smaller groups might easier facilitate in-depth discussions (25), and used an over-recruitment strategy to account for last-minute cancellations.

Data collection

We used semi-structured homogeneous focus groups which were audio recorded and facilitated by MFA (male investigator, certified physiotherapist, and full-time PhD student), who has previous experience in facilitating focus groups, while VS (male investigator, human physiologist, and full-time PhD student) assisted as an observer, took notes, and asked additional questions when necessary.

Neither MFA nor VS had any relations with the stakeholders beforehand. The individual focus groups were held in conference rooms made available by either the stakeholders’ gatekeeper or the researchers. To facilitate group discussions, stakeholders were given three overall statements (Table 1) and instructed to rank them from most to least important and to explain the chosen order. This step was followed by specific questions for individual stakeholder groups to discuss (Table 2). The overall statements were developed based on findings from our previous qualitative study examining the perspectives of clinical and professional experts on the development of community-based exercise programmes for people in antipsychotic treatment (*Andersen et al., submitted*). Only the stakeholders and the researchers (MFA and VS) were present during the focus groups. The focus groups lasted ∼90 minutes and were not repeated. Stakeholders received a token of appreciation of EUR 25.

**Table 1.** Overall statements to facilitate discussion during focus groups.

| **Overall statements** |
| --- |
| *It is important that instructors learn about the treatment of schizophrenia and other psychotic disorders and how the disorders affect people's lives.* |
| *It is important that instructors learn about the concept of recovery and how exercise can support personal recovery.* |
| *It is important that instructors learn about adapting the exercise content, so it is safe and relevant for people in antipsychotic treatment* |

**Table 2.** Specific questions tailored to stakeholders to facilitate discussion in focus groups.

| **Examples of specific questions for stakeholder groups** |  |
| --- | --- |
| Clinical staff from outpatient mental health services | *What can exercise instructors do in order for community-based exercise to complement outpatient mental health services?* |
| Exercise instructors with/without experience in coaching people in antipsychotic treatment | *What concerns do you have about facilitating community-based exercise targeting young adults with schizophrenia?* |
| Physiotherapists working in mental health services. | *What is important to consider when working with community-based exercise targeting young adults with schizophrenia?* |
| Relatives of young adults in antipsychotic treatment | *What do exercise instructors need to be aware of to meet the needs of relatives in relation to community-based exercise targeting young adults with schizophrenia?* |
| Young adults in antipsychotic treatment | *What is important for supervised community-based exercise to facilitate long-term physical active behaviour?* |

Data analysis

A research assistant transcribed the audio recorded material verbatim in accordance with conventions described in a transcription manual. MFA validated the transcripts against the audio recordings. NVivo 12 (QRS International, Melbourne, Australia) and Microsoft Excel were used to assist in data management. All data (in Danish) can be provided upon reasonable request.

Inductive qualitative content analysis, as described by Graneheim and Lundman (26), was applied to analyse the focus group discussions. MFA and KR (female investigator, registered nurse, and full-time PhD student) initially read and reread the transcripts separately before jointly discussing them to obtain a sense of the complete data material. Subsequently, MFA de-contextualised the material, identifying and extracting meaning units before labelling them with descriptive codes. Next MFA compared, abstracted, and sorted codes into categories and subcategories, which were discussed with KR and JM (female investigator, psychologist, senior qualitative health researcher, and clinical professor).

Categories and subcategories were continuously compared with the original transcripts in an iterative analytic process describing the content on a manifest level. To describe the underlying meaning of the categories and subcategories, a single overarching theme was generated on a latent and interpretative level in mutual discussion between MFA, KR and JM.

## Results

### Characteristics of informants

Six focus groups were conducted and comprised a total of 30 individuals representing five different stakeholder groups in January and February 2022. Two focus groups included clinical staff from mental health outpatient services, one of which served as a pilot test of the interview guide. Since the interview guide did not require any revision, the transcript from the pilot focus group was included in the analysis. Table 3 presents the selection criteria, recruitment method and stakeholders’ characteristics.

**Table 3.**
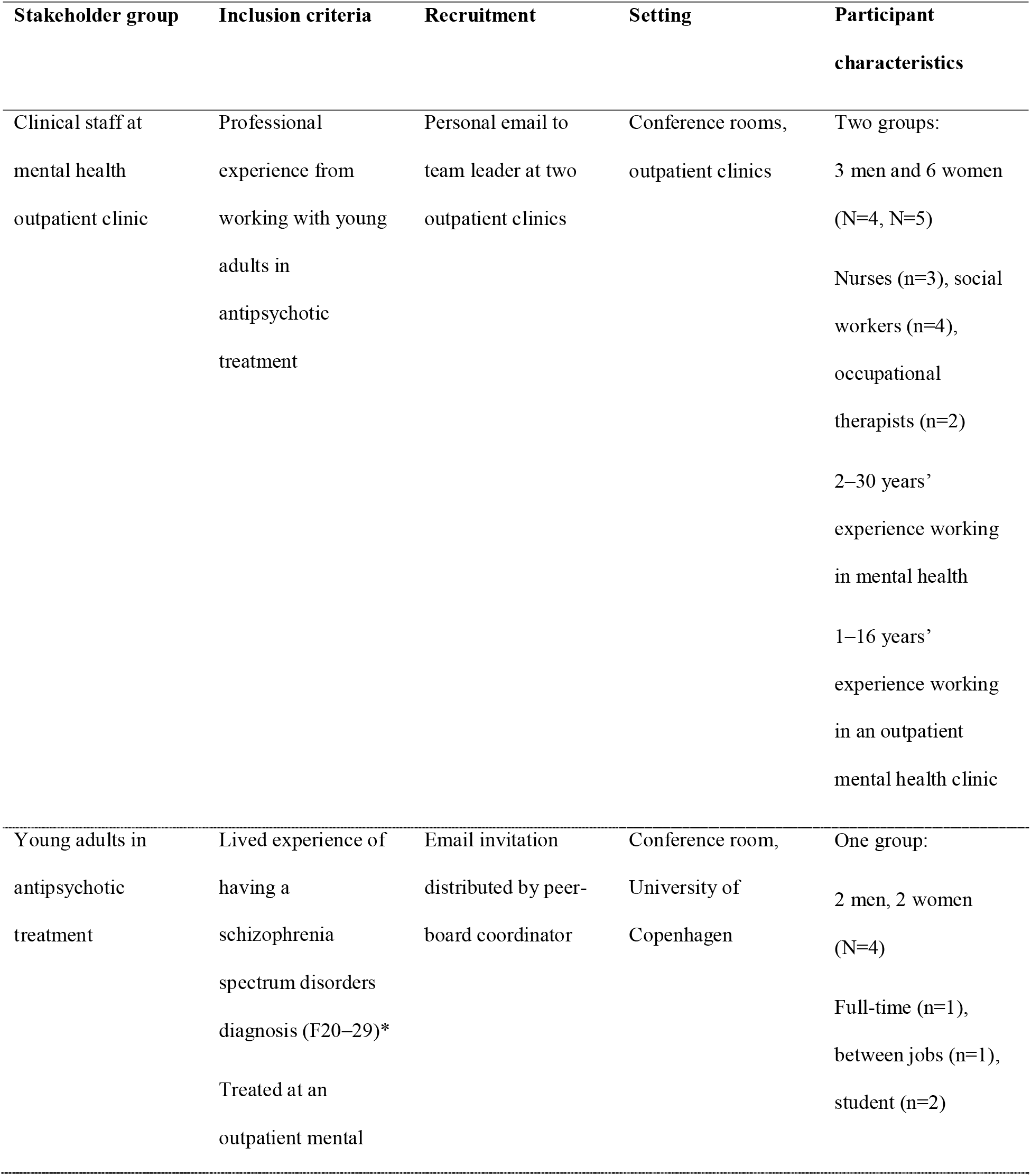
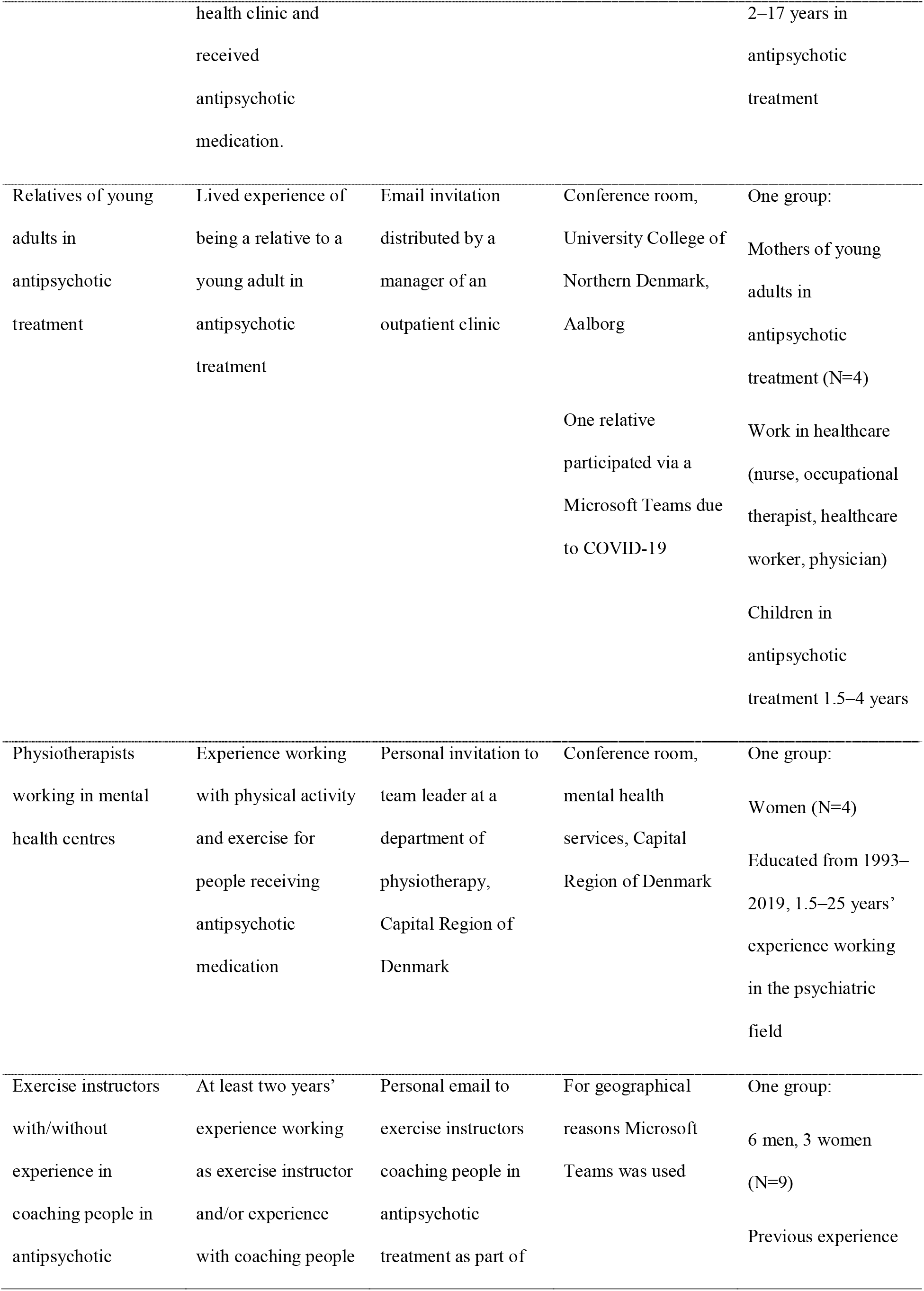
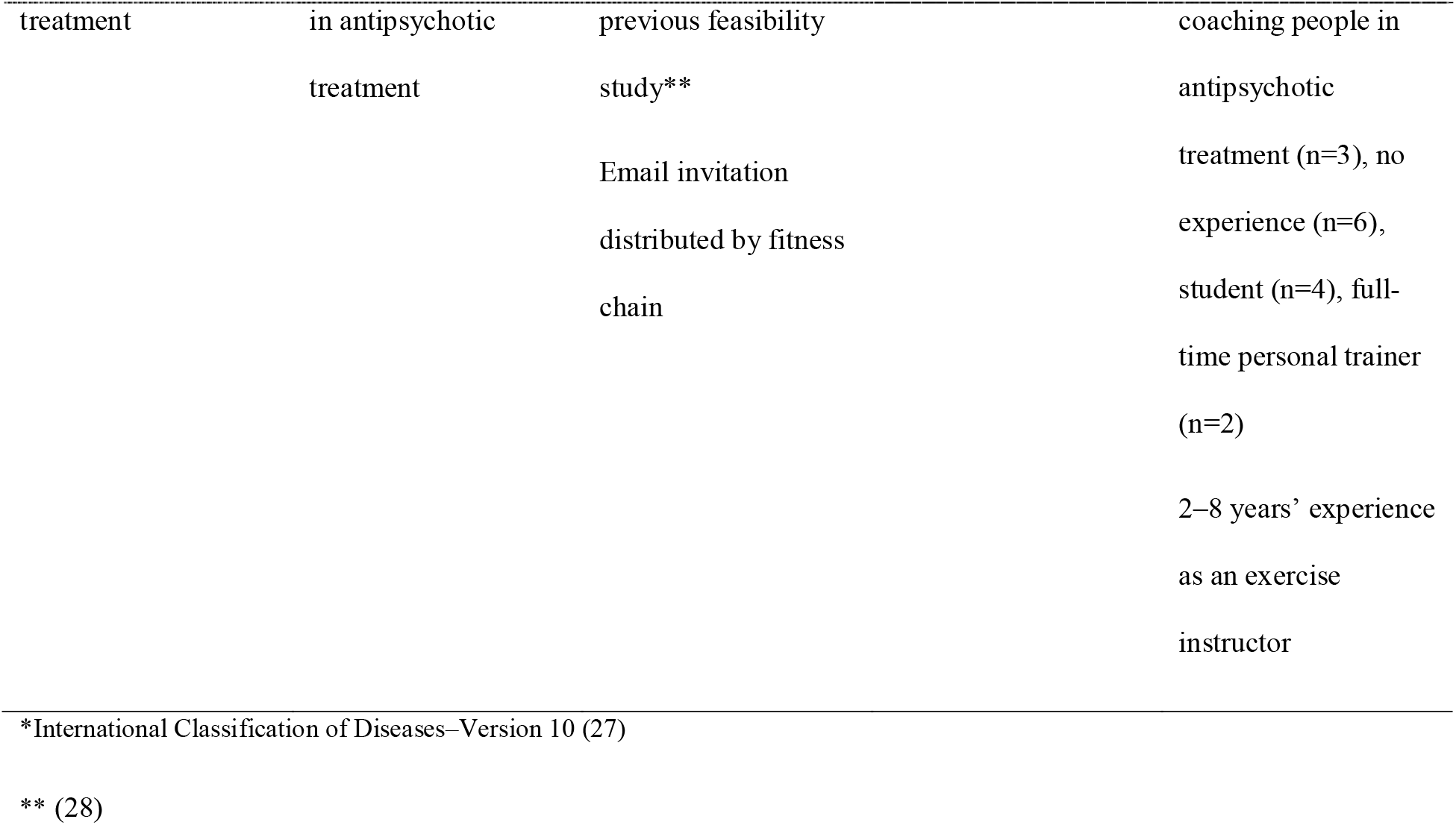
Selection, recruitment, and characteristics of stakeholders (n=30)

### Findings

Thirty-three unique codes, nine subcategories, three categories and one overarching theme were developed (Figure 1). Figure 2 provides an example of the condensation-abstraction process. The three main categories were: *acknowledging mental illness; applying a resource-oriented approach, and promoting exercise as a shared activity,* while the overarching theme was: *instructors as guardians of an inclusive culture*. In the following presentation of the results, non-health professional exercise instructors will be addressed only as *instructors.* Young adults in antipsychotic treatment who participate in community-based exercise will be addressed only as *participants*.

**Figure 1.**
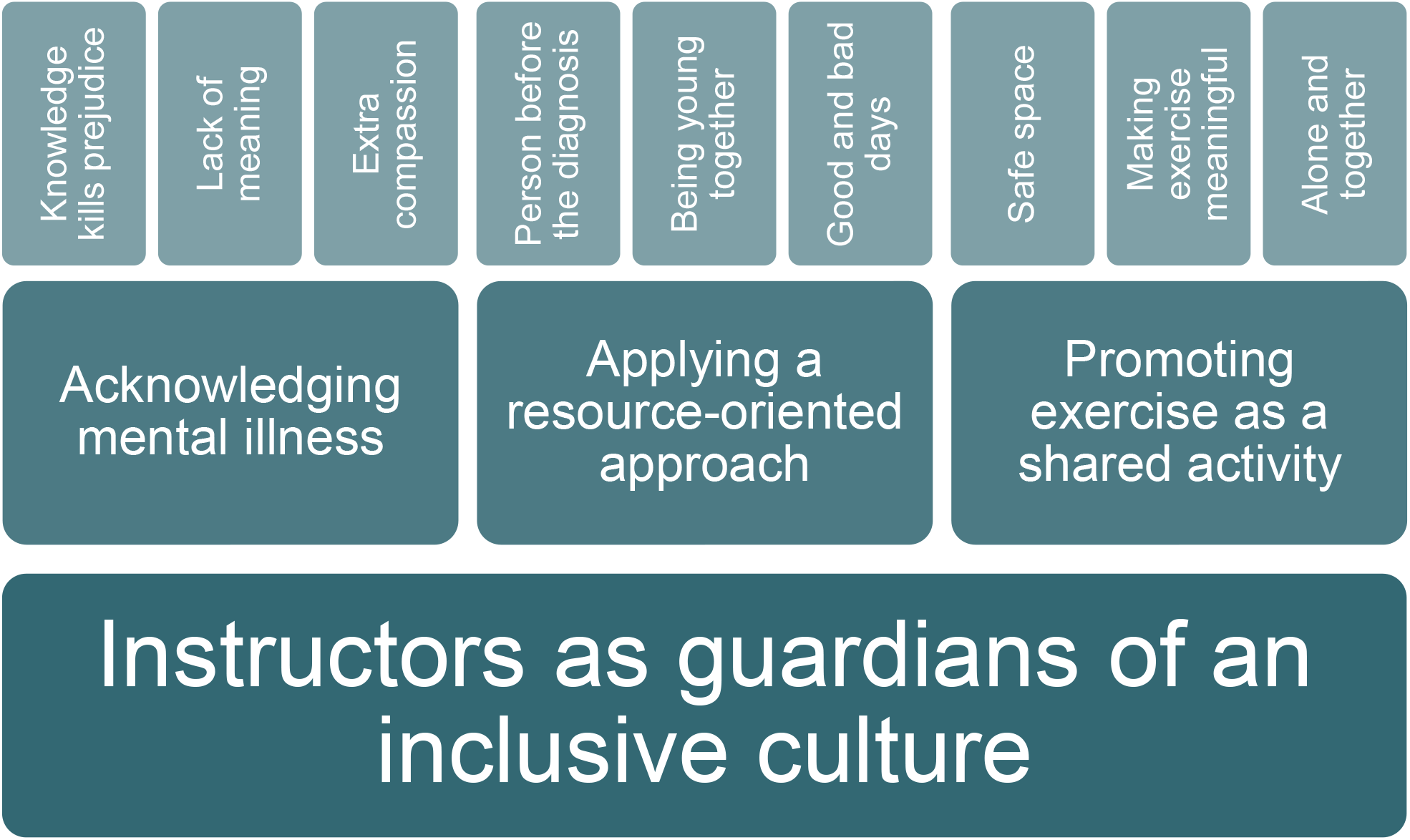
Overview of analysis resulting in nine subcategories, three categories, and one overarching theme.

**Figure 2.**
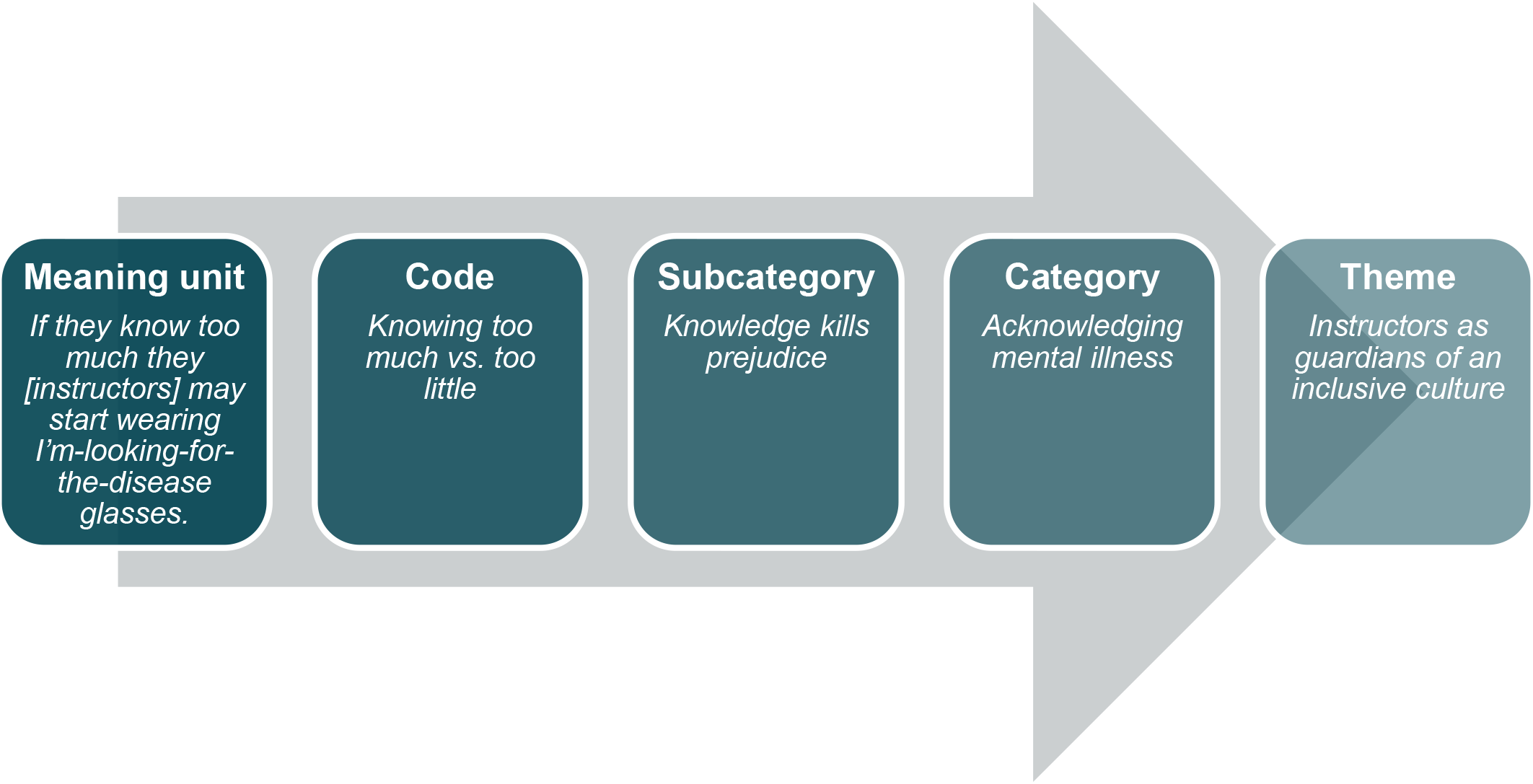
Example of the analytical process from meaning unit to theme.

### Category 1: Acknowledging mental illness

According to the stakeholders, an educational programme should enable instructors to gain an understanding and acknowledgement of what schizophrenia and psychotic disorders are and how the diseases may affect the life of the person suffering from them. Thus, providing instructors knowledge and tools for ongoing reflection, was considered helpful to eliminate common myths and stigma surrounding mental illness.

Based on the claim that *knowledge kills prejudice*, the stakeholders highlighted that instructors should receive basic knowledge about schizophrenia and psychotic disorders. However, knowledge should not be provided with the aim of enabling instructors to engage in therapeutic work. Several stakeholders pointed to the balance between knowing too little or too much, as both may lead to an inappropriate focus on the disease:

> *If they [instructors] know too much they may start wearing “the I’m-looking-for-the-disease glasses”*. (Clinical staff focus group)

The purpose of the knowledge taught should be to reduce stigma by clarifying that limited motivation, interest, and expression are well-known negative symptoms of schizophrenia and potential side effects of antipsychotic treatment, not to be confused with personality traits. Symptoms often fluctuate and are potentially expressed as the feeling that life has a *lack of meaning.* These periods might lead to self-devaluation, a dominant issue discussed among the group of young adults in antipsychotic treatment:

> *It makes me think about the shame. I don’t know if others feel the same way but the feeling of: Why is this so awkward? Why am I making it awkward? Why can’t I figure out how to be in company with other people? I have thought a lot about that.* (Young adult focus group)

At times, participants may be in a vulnerable state of mind, and stressful situations may trigger a fight or flight response. The stakeholders said that instructors may need to show *extra compassion* by acknowledging even the smallest victories and focusing on the effort being made rather than the results achieved. The instructors’ role is to empower the participants and serve as an external motivator during periods when the participant’s internal motivation might be low.

### Category 2: Applying a resource-oriented approach

The stakeholders emphasised that the instructors must meet participants with the same level of curiosity as they would meet anyone else at the gym. They must, however, deliberately focus on exploring the personal resources of the participants.

Thus, an educational programme must inspire instructors to put the *person before the diagnosis* and acknowledge that even though participants may share the same diagnosis, they will all have different difficulties, strengths, preferences, and dreams in life:

> *For me, it’s about being regarded as a person, not just another patient.* (Young adult focus group)

The stakeholders said that instructors must be encouraged to communicate with curiosity if they observe participants who are experiencing challenges due to their mental illness. Psychotic symptoms, such as hallucinations and delusions may inhibit participants from touching exercise equipment, while negative symptoms, such as apathy and withdrawal from social situations may hinder them from participating in a team workout. The stakeholders fear that instructors may refrain from addressing what is happening to avoid making the situation worse, but this may constitute a disservice to the participants:

> *…well, as long as they [instructors] dare ask, they’re not doing any harm. It’s the situations where they don’t ask, that they might miss something*. (Clinical staff focus group)

Instructors should embrace an attitude that shows that they are interested in learning from the participants as experts in their own lives and by building a balanced relationship. Sharing and laughing about everyday problems, thoughts, and experiences may provide an atmosphere where being physically active with others is a natural part of *being young together*. Thus, the exercise community could serve as a distraction from the disease and a place for people to step out of the role as a patient. Furthermore, doing what other young adults are doing may help boost the participants’ identity of being young and fitting in. This was something especially the relatives of the young adults in antipsychotic treatment emphasised:

> *So, she [a young adult in antipsychotic treatment] can take a selfie for her friends and tag it with “going to the gym, smiley” helping her maintain an identity similar to her friends.* (Relatives focus group)

To enable participants to be physically active despite potential barriers, stakeholders highlight that an exercise community should be able to embrace participants during *good and bad days*:

> *Once, I heard on the radio that you can climb a hill in different ways. You can run, you can walk, and you can crawl…but we all need to get up there.* (Young adult focus group)

An educational programme should ideally give instructors the ability to balance the demands of the exercise content with the physical, psychological, and social capacity of the participants while acknowledging when to take care of special needs and when to push participants out of their comfort zones. However, the stakeholders recommended starting slowly to improve the chance of success, which is why they may benefit from discreetly making note of the energy level of the participants at the beginning of an exercise session.

### Category 3: Promoting exercise as a shared activity

According to stakeholders, a shared exercise community may become a symbol of the relationship between the participants, their peers, and the instructors which may in turn produce personal growth for everybody involved. Nevertheless, the instructors are responsible for contextual factors that may be crucial to realising the potential of exercising together.

As such, community-based exercise should represent a *safe space* for participants that ensures a recognisable and predictable structure and the presence of trusted others. The stakeholders also mentioned that instructors should be adequately prepared for their role as instructors for them to feel comfortable. Instructors will likely be affected if they see signs of self-harm, such as cutting marks and cigarette burns, but their focus should be to support participants in their recovery process and not to solve their difficulties:

> *Trust that others [health professionals] have the responsibility to address this, not themselves [instructors]. They are just one part of the journey for the young person to feel better. No further demands should be imposed on the instructors besides doing the best they can and knowing that they are doing something good.* (Relatives focus group)

To facilitate empowerment and sustained engagement in physical activity, the stakeholders emphasized the importance of *making exercise meaningful* for everyone, even though this may be challenging when the exercise is delivered in groups. Aligning expectations may be helpful in identifying individual goals, and instructors can help support minor improvements according to these goals. For some, this may involve assisting with providing structure in everyday life; for others, it may involve providing an arena for participants to develop their social skills. Some participants may strive to be strong or fit, while others might seek a distraction from clinical symptoms. However, the stakeholders believe that the overall goal of the exercise should be that it is fun. One exercise instructor with experience in coaching young people in antipsychotic treatment stated:

> *Having a playful part at the beginning [of an exercise class] really loosened up the atmosphere…playing around positively affected the dynamic…and ending the class well…having a good experience to go home with.* (Exercise instructor focus group)

Community-based exercise should provide space for being both *alone and together* and focus on both individual and joint goals. Facilitation of group dynamics is crucial since doing so produces a positive, binding community that can support continuous participation. Exercising with others, who might experience some of the same challenges due to mental illness may produce a mutual understanding which does not require to be articulated. As such, instructors should emphasise community-based exercise as a joint space that participants share in (re)building their exercise identity. In other words, community-based exercise can act as a steppingstone towards social inclusion in civil life:

> *It can be a steppingstone to something else. It’s a pleasant thought because we sometimes lack this kind of steppingstone. I think it’s cool…and cool it doesn’t “smell like psychiatry”.* (Clinical staff focus group)

### Overarching theme: Instructors as guardians of an inclusive culture

An overall objective of an educational programme is to give the instructors the opportunity to build a role as *guardians of an inclusive culture.* A culture that has a clear anti-stigma approach to people with mental illness and ensure achievable physical activity for everyone regardless of physical, social or mental barriers. As such, their focus should be on identifying personal resources in each participant rather than limitations resulting from mental illness. Further, instructors should be aware that their attitude and behaviour might be the subject of attention and that participants may admire and be inspired by them. The instructors should draw attention away from themselves and invite participants to embrace the protective nature of the exercise community, in addition to being part of forming it. Lastly, instructors should be prepared to take responsibility for ensuring a motivating exercise environment, delivering exercise designed to improve the participants’ feeling of being strong and fit and thus provide the participants an opportunity to (re)build an exercise identity.

## Discussion

This study aimed to provide perspectives into the required elements of an educational programme for non-health professional exercise instructors delivering community-based exercise targeting young adults with schizophrenia, we attempt to fulfil the increasing request for sustainable, easily accessible, and engaging community-based initiatives in mental health care (30). Overall, our findings suggest that educational programmes should allow instructors to promote a culture that embraces an anti-stigma stance by: a) acknowledging the difficulties that mental illness may produce; b) taking a resource-oriented approach to the participants; and c) using exercise as a shared activity that symbolises the relationship between participants, their peers, and the instructor. Notably, stakeholders emphasise the highly significant role of instructors in producing and protecting an inclusive culture. A qualitative review of physical activity among people with enduring mental health difficulties found that exercise instructors were essential in providing a safe and supportive environment while bolstering their sense of competence and self-esteem (31). Furthermore, our previous qualitative study on the experiences of participants and instructors investigating the feasibility of an exercise programme for people with first-episode psychosis indicated that non-health professional exercise instructors contributed considerably to a caring yet challenging environment (32). Instructors may also play an important role in creating a feeling of partaking in normal physical activity with other young adults, helping to reverse the negative stigma associated with mental illness (10).

The stakeholder groups agreed that exercise instructors should receive training in basic mental health literacy, which is supported by an international consensus statement (29). Especially knowledge about negative symptoms as these may often be misinterpreted as personal traits and lead to stigma towards people with mental illness among the general population. Furthermore, negative symptoms are highly associated with social functioning and self-efficacy (33) and thereby strongly affect participants’ ability to participate in community-based exercise. Another notable finding across the stakeholders was the importance of the community and that an educational programme should provide the instructors with the skills to facilitate social connectivity among the participants, which is supported by current evidence (10). Qualitative findings show that the main narratives related to participation in sports and exercise were that the experience of achievement was shared by, and could potentially be shared with, many others in everyday life (34). Instructors must nevertheless be aware that social interaction is difficult for most people with schizophrenia and psychotic disorders and may lead to self-isolation (35). Gym-based group exercise may provide unique flexibility that allows participants to oscillate between socialising and exercising alone (32). Thus, instructors should be able to scale exercise content on physical, social and mental parameters in order to make exercise meaningful for each participant in the exercise community. Indeed, the motivation for being physically active varies greatly among people with schizophrenia. Such motivational factors may range from physical outcomes, e.g., weight control and fitness, to more psychological and social outcomes, e.g., improving mood and meeting new people (12,36).

The stakeholders pointed to the importance of balancing knowing too much and too little. Similarly, the educational content should allow instructors to balance between promoting exercise in a non-patient environment while respecting the limitations and boundaries of people in antipsychotic treatment. Indeed, based on the assumption that knowledge should serve to promote anti-stigma, education appears to be effective in terms of improving knowledge and positive attitudes toward reducing stigma and discrimination regarding mental illness (37). Furthermore, an educational programme should facilitate ongoing reflections among the instructors in order to obtain practical knowledge. Qualitative evidence suggests the importance of exchanging experiences and ideas with health professionals and peer instructors when non-health professionals supervise physical activity in clinical populations (38). Also of importance, exercise instructors may represent an exercise evangelistic mindset assuming that exercise participation will always contribute to positive personal growth and mental well-being. Thus, education should clarify that, besides the potential benefits of exercise and sports, it also bears the risk to do harm, e.g., due to social exclusion or comparison to more successful peers (10). This is especially important bearing in mind, that initiating community-based exercise in this population may be associated with low self-esteem and the feeling of powerlessness after previously failed attempts to initiate physical activity (7,9).

### Methodological considerations

Some limitations and strengths should be considered. A strength is that we recruited individuals from five different stakeholder groups, all highly relevant and with specific knowledge and experience relevant to the objective of the current study. However, due to the COVID-19 pandemic, numerous stakeholders cancelled at the last minute. Consequently, a possible limitation is that only four people from the focus group for young adults in antipsychotic treatment and four from the focus group for relatives participated and that they may be characterized by having stronger resources in comparison to people who were not able or willing to participate. Another limitation of relevance to all groups is that the sampling strategy potentially led to the recruitment of stakeholders with a special interest in, and a predominantly positive attitude towards, exercise. In this regard, relatives were also only represented by mothers working in the healthcare sector, which may challenge the transferability of their perspectives to other relatives, e.g., fathers and relatives not working in the healthcare sector. Using inductive content analysis allowed us to capture the study’s intended objective, which has been underexplored by previous research. Our findings present an overall theme that encompasses a comprehensive interpretation of stakeholder perspectives; hence, we acknowledge the influence of our own preconceptions (39). To enhance credibility, continuous reflections and discussions on preconceptions relating to the study objective were carried out throughout the entire research process (40).

### Implication for practice

The results of the current qualitative study investigating the perspectives of different stakeholder groups contributed to the development of an educational programme for non-health professional exercise instructors delivering community-based exercise targeting young adults in antipsychotic treatment currently implemented as a part of the Vega trial (ref). The educational programme encompasses 1) a written exercise instructor manual; 2) a one-day educational course; and 3) the possibility of a continuous exchange of experiences between instructors. Appendix 2 provides an overview of the educational programme with headlines of the educational content. The full version of the educational programme in Danish can be provided upon reasonable request.

## Conclusion

An educational programme for exercise instructors delivering community-based exercise to young adults in antipsychotic treatment should include educational content that enables instructors to promote exercise as a shared activity while acknowledging mental illness and having a resource-oriented approach. Indeed, instructors must act as guardians of an inclusive culture. Our results may be transferable to the education of non-health professional exercise instructors in other areas of mental health care where the aim is to facilitate sustainable, recreational, community-based activities.

## Declarations

### Ethical approval and consent to participate

In addition to receiving written and oral information about the nature of the study prior to participation, all participants provided written informed consent and were guaranteed anonymity and confidentiality. The study was conducted in accordance with the Declaration of Helsinki. The Regional Ethics Committee of Northern Denmark has confirmed that no formal ethical approval was required (Case No. 2023000206) for the current study.

## Availability of data and materials

Data and material can be provided upon reasonable request by the corresponding author.

## Authors’ contributions

All authors participated in conceptualizing the study. MFA, KR and JM planned the study design and methodology. MFA and VS were responsible for the data collection and MFA was responsible for the initial data analysis. MFA, KR and JM contributed to the data analysis process. MFA generated the first draft, and all authors critically revisited the draft for important intellectual content. Lastly, the final version was sent to all authors for approval. All authors read and approved the final manuscript.

## Data Availability

All data produced in the present study are available upon reasonable request to the authors

## Acknowledgements

The authors are thankful to the stakeholders who participated in this study, for giving their time and providing detailed accounts of their perspectives to make this work possible. Furthermore, the authors are thankful to Anna Jessen (research assistant) for assisting in the transcription process and Mads Juul Christensen (PhD student) for assisting as an observer in the focus group with relatives of young adults in antipsychotic treatment.

